# Mitochondrial fatty acid synthesis is essential for coordinated energy transformation

**DOI:** 10.1101/2023.04.03.23288010

**Authors:** Friederike Haumann, Ioannis Evangelakos, Anna Worthmann, Imke Liebold, Stefan Kotschi, Almut Turid Bischoff, Christiane M. Neuhofer, Michaela Schweizer, Markus Heine, the mitoNET consortium, Boriana Büchner, Thomas Klopstock, Cornelia Prehn, Kenneth Allen Dyar, Holger Prokisch, Lidia Bosurgi, Joerg Heeren, Alexander Bartelt, Christian Kubisch, Christian Schlein

**Affiliations:** Institute of Human Genetics, University Medical Center Hamburg-Eppendorf, Germany; Department of Biochemistry and Molecular Cell Biology, University Medical Center Hamburg-Eppendorf, Germany; I. Department of Medicine, University Medical Center Hamburg-Eppendorf, Germany; Protozoa Immunology, Bernhard Nocht Institute for Tropical Medicine, Hamburg, Germany; Institute for Cardiovascular Prevention (IPEK), Ludwig-Maximilians-University of Munich, Munich, Germany; Department of Neurology, Friedrich-Baur-Institute, University Hospital LMU Munich, Munich, Germany; Technical University of Munich, School of Medicine, Institute of Human Genetics, Munich, Germany; Institute of Neurogenomics, Helmholtz Zentrum Munich, Munich, Germany; Core Facility of Electron Microscopy, Center for Molecular Neurobiology ZMNH, University Medical Center Hamburg-Eppendorf, Hamburg, Germany; Metabolomics and Proteomics Core Facility, Helmholtz Zentrum Munich, German Research Center for Environmental Health (GmbH), 85764 Neuherberg, Germany; Institute for Diabetes and Cancer, Helmholtz Zentrum Munich, German Research Center for Environmental Health, 85764 Neuherberg, Germany; German Center for Diabetes Research (DZD), 85764 Neuherberg, Germany; German Center for Cardiovascular Research, Partner Site Munich Heart Alliance, LMU Hospital, Munich, Germany; Department of Molecular Metabolism & Sabri Ülker Center for Metabolic Research, Harvard T.H. Chan School of Public Health, Boston, USA

## Abstract

Mitochondria warrant cellular energy demands by generating energy equivalents in central carbon metabolism. They are also able to newly synthesize fatty acids via mitochondrial fatty acid synthesis (mtFAS), however, the role of mtFAS for systemic metabolism has been poorly investigated. Here we show that mitochondrial Trans-2-Enoyl-CoA Reductase (MECR), a key enzyme of mtFAS, critically regulates cellular and systemic glucose and lipid homeostasis. In mice, liver or adipose tissue-specific deletion of *Mecr* reduces the capacity for aerobic glycolytic catabolism and lipogenesis and causes severe mitochondrial as well as fatal parenchymal organ dysfunction. Mechanistically, mtFAS is essential for pyruvate dehydrogenase activity, resulting in low NAD(P)H synthesis and reduced non-mitochondrial lipogenesis. In different human mitochondriopathies we further identify a dysregulation of mtFAS-associated lipid species, thus linking inherited mitochondrial disease to mtFAS. In summary, we introduce mtFAS as an important player in metabolic health via facilitating cellular glycolysis-derived metabolite transformation ultimately linking mtFAS to mitochondrial function and diseases.

## Introduction

Mitochondria are key organelles of catabolic metabolism and produce energy equivalents from lipid or carbohydrate degradation products (Spinelli and Haigis, 2018). However, mitochondria are also anabolic organelles as they are capable to synthesize hormones (Papadopoulos and Miller, 2012), release carnitine-bound fatty acids as well as they are able to newly synthesize fatty acids (Hiltunen et al., 2009). Mitochondrial fatty acid synthesis (mtFAS) is an evolutionary ancient mechanism that is very similar to fatty acid synthesis in bacteria and duplicates the cytosolic fatty acid synthesis pathway (Nowinski et al., 2018). This canonical *de novo* lipogenesis pathway takes place in the cytosol and generates palmitate via the multi-enzymatic fatty acid synthase (FASN) protein. In contrast, in mtFAS the different steps of fatty acid synthesis are carried out by many single enzymes within the mitochondrial matrix. One of these enzymes is the mitochondrial trans-2-enoyl-CoA reductase encoded by *MECR*, which is known to catalyze the last step of mtFAS, producing a medium and long chain acyl-residue bound to the acyl-carrier protein (ACP, encoded by *NDUFAB*) in mitochondria (Tanvir Rahman et al., 2023). In 2016, biallelic *MECR* mutations were firstly reported to cause a putative neurodegenerative syndrome, in which the patients mainly suffer from optic nerve atrophy and muscular dystonia (Heimer et al., 2016). Whereas the role of cytosolic fatty acid synthesis is well described and is a topic of interest for therapeutically targeting non-alcoholic steatohepatitis (Loomba et al., 2021; Worthmann et al., 2022) and cancer (Falchook et al., 2021), the role of mtFAS in systemic lipid and glucose metabolism is less characterized.

*In vitro* experiments revealed that mtFAS is required for efficient synthesis of lipoic acid, a cofactor of pyruvate dehydrogenase (PDH) and as such, an important mediator of glucose degradation (Nowinski et al., 2018). Moreover, lipoic acid acts as a cofactor for alpha-ketoglutarate synthase, 2-oxoadipate dehydrogenase, branched-chain ketoacid dehydrogenase, and the glycine cleavage system (Mayr et al., 2014) and reflects an important redox factor (Solmonson and DeBerardinis, 2018). In humans, in addition to *MECR* also other genes are known to affect lipoic acid synthesis and usage, such as *LIAS* (Baker et al., 2014; Mayr et al., 2011), *DLD* (Elpeleg et al., 1997), *LIPT2* (Habarou et al., 2017), *BOLA3* (Baker et al., 2014; Cameron et al., 2011; Haack et al., 2013; Saudino et al., 2021), *NFU1* (Cameron et al., 2011; Navarro-Sastre et al., 2011), *GLRX5* (Baker et al., 2014; Camaschella et al., 2007), *IBA57* (Ajit Bolar et al., 2013) or *MCAT* (Li et al., 2020); however, the whole body impact varies largely from (MCAT-associated-) Leber’s hereditary optic neuropathy (LHON)-like disease to early death by cardiac failure (*BOLA3*-associated), indicating a cell type- and organ-specific role of the different enzymes. Of note, studies in mice showed that supplementation of exogenous lipoic acid was not able to fully rescue defects of endogenous lipoic acid synthesis, suggesting a lack of transport mechanisms of lipoic acid into mitochondria (Tajima et al., 2019). Nevertheless, it has been implied that supplementation of lipoic acid might have anti-diabetic effects (Salehi et al., 2019), indicating that also extramitochondrial lipoic acid supplementation might have a beneficial effect, e.g. via increasing anti-oxidant capacities. Of note, the function of mtFAS for systemic lipid and glucose metabolism has not been explored and, thus, its putative role for the development and progression of metabolic disease such as obesity, type 2 diabetes mellitus and dyslipidemia is unknown.

Here, we identify *MECR* as an important modulator of glucose and lipid homeostasis. We show that tissue specific deletion of *Mecr* in adipose tissue or liver in mice disturbs systemic glucose and lipid disposal and energy metabolism. While adipocyte-specific *Mecr* loss induces adipose tissue dysfunction which is characterized by metabolic stress, adipose tissue inflammation, reduced glucose and lipid tolerance as well as impaired thermogenesis in brown fat, liver-specific deletion of *Mecr* causes hepatic megamitochondria formation, ultimately resulting in failure to thrive and early death. Mechanistically, we show that MECR reduces protein lipoylation and critically determines PDH activity and thus availability of cellular energy equivalents. Human mtFAS dysfunction rearranges cellular glucose metabolism, which might also affect the outcome of other mitochondriopathies. In summary, we identify mtFAS as a pivotal regulator of metabolic health as it mediates the capability of functional energy dissipation of mitochondria.

## Results

### Loss of adipocyte *Mecr* leads to impaired energy breakdown and insulin resistance

Adipose tissues are the major sources of *Mecr* expression (Fig.S1a). To characterize the impact of *Mecr* deficiency in adipose tissues, we generated *Mecr*^*flox/flox*^ *Adipoq-Cre* mice. MECR is described to be involved in lipoic acid synthesis in vitro (Nowinski et al., 2018) and indeed, in white adipose tissue, loss of MECR leads to decreased protein lipoylation (Fig.1a), but also to reduced succinylation, indicating disturbed mitochondrial succinate metabolism. Lipoic acid is a critical cofactor for PDH activity, which was decreased in adipose tissue (Fig.1b). Of note, PDH E1 subunit alpha 1 (PDHA1), mirroring total PDH level, was increased on protein level and its inhibitory phosphorylation was nearly absent (Fig.1a), indicating compensatory effects to restore the attenuated PDH activity. Further, electron micrographs of WAT showed a disturbed mitochondrial structure in the *Adipoq Cre*^*+*^ mice characterized by reduced cristae structure and a cigar-like shaped, electron-dense mitochondrial form (Fig.1c). To analyze any impact on the lipidomic signature of the mice, we performed lipidomic analysis of plasma, white and brown adipose tissues as well as liver. Except for slight reductions in DAG/TAG, the lipidome of plasma and liver remained mostly unaffected by adipocyte-specific loss of *Mecr*. In contrast, in brown and even more pronounced in white adipose tissue all lipid classes were reduced in *Adipoq-Cre*^+^ mice compared to the respective organ of the *Adipoq-Cre*^-^ littermates (control), emphasizing a general cellular lipid metabolism dysfunction (Fig.1d). We further applied a targeted metabolomics approach to analyze other potentially changed metabolites. Here, we additionally identified changes in amino acids, and sulfonic acids – in particular taurine (FigS1b) – indicating that MECR affects various metabolites and might be involved in many metabolic pathways in WAT or that the cells adapt by rearrangement of metabolic pathways (Fig.1e). Gene expression analysis revealed a considerable increase of cytosolic lipid synthesis-related genes in WAT upon *Mecr* deficiency as well as slightly elevated markers of fibrosis (*Timp1, Cxcl10*) and cell stress (*Ddit3, Ccl2*) (Fig.1f). However, this was not accompanied by an inflammatory cell infiltration, indicated by neither altered inflammatory markers (*Tnf, Cd68*) on gene expression level (Fig.1f) nor innate/adaptive immune cell population-enrichement in the WAT of the *Adipoq-Cre*^*+*^ mice(Fig.3g). Similarly, among the “metabolically active macrophages” defined as CD45^+^Ly6G^-^CD11b^+^F4/80^+^CD11c^+^ (Hill et al., 2018; Jaitin et al., 2019; Shaul et al., 2010), no differences in the expression of markers which better define their identity (CD68 and Ly6C) as well as their polarization status (ARG1, CCR2, CD80 and CX3CR1) were observed between *Adipoq-Cre*^*+*^ and *Adipoq-Cre*^*-*^ mice. To further investigate the ability of functional cellular lipid metabolism of WAT cells, we performed *in vitro* lipolysis assays in primary adipocytes. This showed that both basal as well as norepinephrine-induced lipolysis and free fatty acid release were increased in primary adipocytes derived from *Adipoq-Cre*^*+*^ mice (Fig.1i). At the same time, while the release of free fatty acids could be suppressed by insulin in *Adipoq*-*Cre*^*-*^ adipocytes, this effect was attenuated in *Adipoq*-*Cre*^*+*^ adipocytes (Fig.1j) indicating cellular insulin resistance. This was also accompanied by reduced phospho-AKT levels, a surrogate marker of impaired insulin signaling (Fig.S1c). In line, *in vivo* VLDL production, which is highly dependent on the amount of free fatty acids released from white adipose tissue, was higher in *Adipoq-Cre*^*+*^ mice (Fig.1k). To further estimate the role of *Mecr* for the development of insulin resistance, we fed *Adipoq-Cre* mice with a high fat diet for 12 weeks and performed a combined radioactive tracer-spiked oral glucose and fat tolerance test. Interestingly, despite the fact that *Adipoq-*Cre^*+*^ mice similarly gained body weight (Fig.S1d), they exhibited a decreased glucose and lipid tolerance (Fig.1l-m), which might be explained by lower uptake of ^14^C-triolein tracer in WAT (Fig.1n) and the reduced PDH activity in obese WAT (Fig.1o) needed for efficient glucose breakdown.

**Figure 1:**
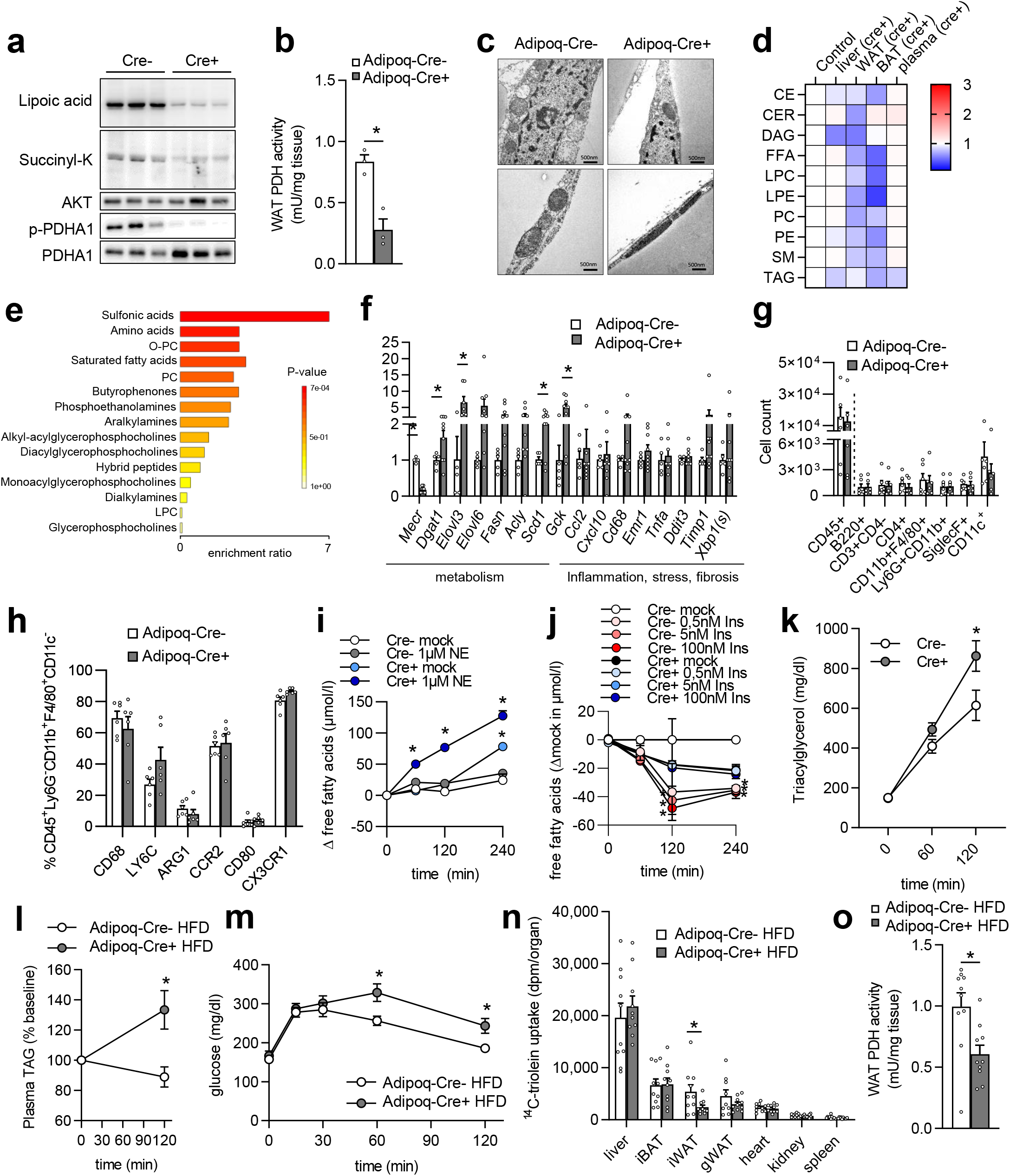
Loss of adipocyte Mecr leads to impaired energy breakdown and insulin resistance. (a) Western blot (n=3), (b) WAT PDH activity (n=3), (c) electron microscopy (n=3) of white adipose tissue from *Mecr*^*flox/flox*^ *Adipoq-Cre*^*+*^ mice and *Cre*^-^ littermate controls. (d) Lipidomics of liver, white adipose tissue (WAT), brown adipose tissue (BAT) or plasma from *Mecr*^*flox/flox*^ *Adipoq-Cre* mice (n=4). (e) Enrichment plot of targeted metabolomics (n=6) from gonadal white adipose tissue of *Mecr*^*flox/flox*^ *Adipoq-Cre* mice. (f) Gene expression (n=5-9) and (g-h) flow cytometry analysis (n=6) of white adipose tissue of *Mecr*^*flox/flox*^ *Adipoq-Cre* mice. (i) Primary adipocytes were differentiated for 10 days and were treated with norepinephrine or mock (n=4) and (j) insulin or mock (n=4) for the indicated time period and free fatty acids as a marker of lipolysis was measured from cell culture supernatant. (k) VLDL synthesis experiment in *Mecr*^*flox/flox*^ *Adipoq-Cre* mice by injection of lipase inhibitor tyloxapol and measurement of the accumulating triacylglycerols in plasma in *Mecr*^*flox/flox*^ *Adipoq-Cre* mice during fasting over time. (l-o) *Mecr*^*flox/flox*^ *Adipoq-Cre* mice were fed a high fat diet for 12 weeks and were gavaged with a combined radioactive tracer-spiked (l) lipid and (m) glucose load where (n) ^14^C-triolein uptake as well as (o) postprandial WAT PDH activity was measured (n=9-10). Error bars indicate standard error of the mean (SEM). *= p<0.05 by two-way ANOVA and Fishers-LSD Test (i, j) or Student’s T-Test.

### Loss of adipocyte *Mecr* leads to brown adipose tissue inflammation and dysfunctional thermogenesis

Compared to other organs and most likely due to its high mitochondrial density, brown adipose tissue (BAT) exhibits the highest gene expression of *Mecr* (Fig.S1a). Of note, when mice are exposed to cold conditions (7 days at 4°C ambient temperature), gene expression levels of mtFAS genes (*Mcat, Mecr, Oxsm, Ndufab1, Cbr4*) increased in BAT (Fig.S2a), suggesting a role during cold adaptation. Besides the drop in most membrane lipid classes (shown in Fig.1d), targeted metabolomic analysis indicated severe changes in amino acid and biotin metabolism in BAT of *Adipoq-Cre*^*+*^ mice (Fig.2a; Fig.S2b). Furthermore, similarly as observed in WAT (Fig.1a), BAT tissue showed that loss of *Mecr* strongly diminished protein lipoylation and the inhibitory PDHA1 phosphorylation (Fig.2b), again suggesting compensatory loss of PDH inhibiting/inactivating mechanisms. In addition, mitochondrial morphology was highly affected in BAT of *Adipoq-Cre*^*+*^ mice. Electron micrographs revealed a different ultrastructure and reduced mitochondria to ER contact sites (marked in green), while ER-lipid droplet contacts (marked in red) appeared to be enriched (Fig.2c). Based on these observations, we hypothesized that glucose catabolism and uptake as well as lipid metabolism would be disturbed in *Adipoq-Cre*^*+*^ mice. Indeed, when we performed radioactive tracer studies, BAT uptake of ^3^H-deoxyglucose (DOG) (Fig.1d) and ^14^C-triolein (Fig.1e) was reduced in *Adipoq-Cre*^+^ mice on chow diet at conventional housing temperature, suggesting lower activity of the tissue or a diminished capacity to handle nutrients for thermogenesis. To evaluate thermogenic capacity, we performed indirect calorimetry experiments. When injected with the beta3-adrenergic agonist CL316,243 to specifically activate thermogenic adipose tissues, *Adipoq-*Cre^*+*^ mice exhibited lower energy expenditure and oxygen consumption (Fig.1f, Fig.S2c,d), indicative of mitigated thermogenic function. When we housed the mice at cold conditions (4°C), *Adipoq-*Cre^*+*^ mice appeared to be cold intolerant as to our surprise their survival was critically reduced (Fig.2g) and the experiment had to be terminated 3 days after the initiation of cold exposure. Cold-exposed BAT of the *Adipoq-Cre*^*+*^ mice presented lower protein levels of UCP1 and OXPHOS complexes (Fig.1h), indicating that impaired non-shivering thermogenesis and mitochondrial respiration might be culpable for their lower survival. Gene expression analysis of BAT at conventional housing temperature showed that genes related to cytosolic lipid synthesis were downregulated while several inflammatory and stress markers were markedly induced. Interestingly, an almost 25-fold increase was noted for the fibrosis marker *Timp1* (Fig.1i). To evaluate whether the inflammation was attributed to brown but not white adipocytes, similar gene expression analysis was performed using *Ucp1-Cre*^*+*^ mice, which resembled the phenotype of *Adipoq-Cre*^*+*^ (Fig.S3a-d). Flow cytometry analysis of BAT showed largely elevated cell counts of several immune cell populations in the *Adipoq-Cre*^*+*^ mice, including CD4+ cells and macrophages (CD11b^+^F4/80^+^CD11c^+^ and CD11b^+^F4/80^+^CD11c^-^) eosinophils (SiglecF^+^). (Fig.2j). Further phenotypical characterization of a population of metabolically active macrophages (CD11b^+^F4/80^+^CD11c^+^) revealed differences in the frequency of cells expressing CD68 and Ly6C antigens as well as alteration in their polarization status with high levels of molecules associated to a tissue remodeling/pro fibrotic phenotype (CX3CR1 and ARG-1) and antigen presentation capacity (CD80) in *Adipoq-Cre*^*+*^ mice compared to wild type littermates (Fig.2k). Of note, the increased number of non-adipocytes in BAT of *Adipoq-Cre*^*+*^ mice might explain some of the differences in lipidomic and metabolomic analysis due to the massive infiltration of immune cells and a dilution of brown adipocyte analytes. In sum, both white and brown adipose tissue of mice lacking *Mecr* in adipocytes showed a severe parenchymal dysfunction, which was accompanied by higher markers of inflammation and fibrosis in BAT, indicating that mtFAS is essential for regular adipose tissue function.

**Figure 2:**
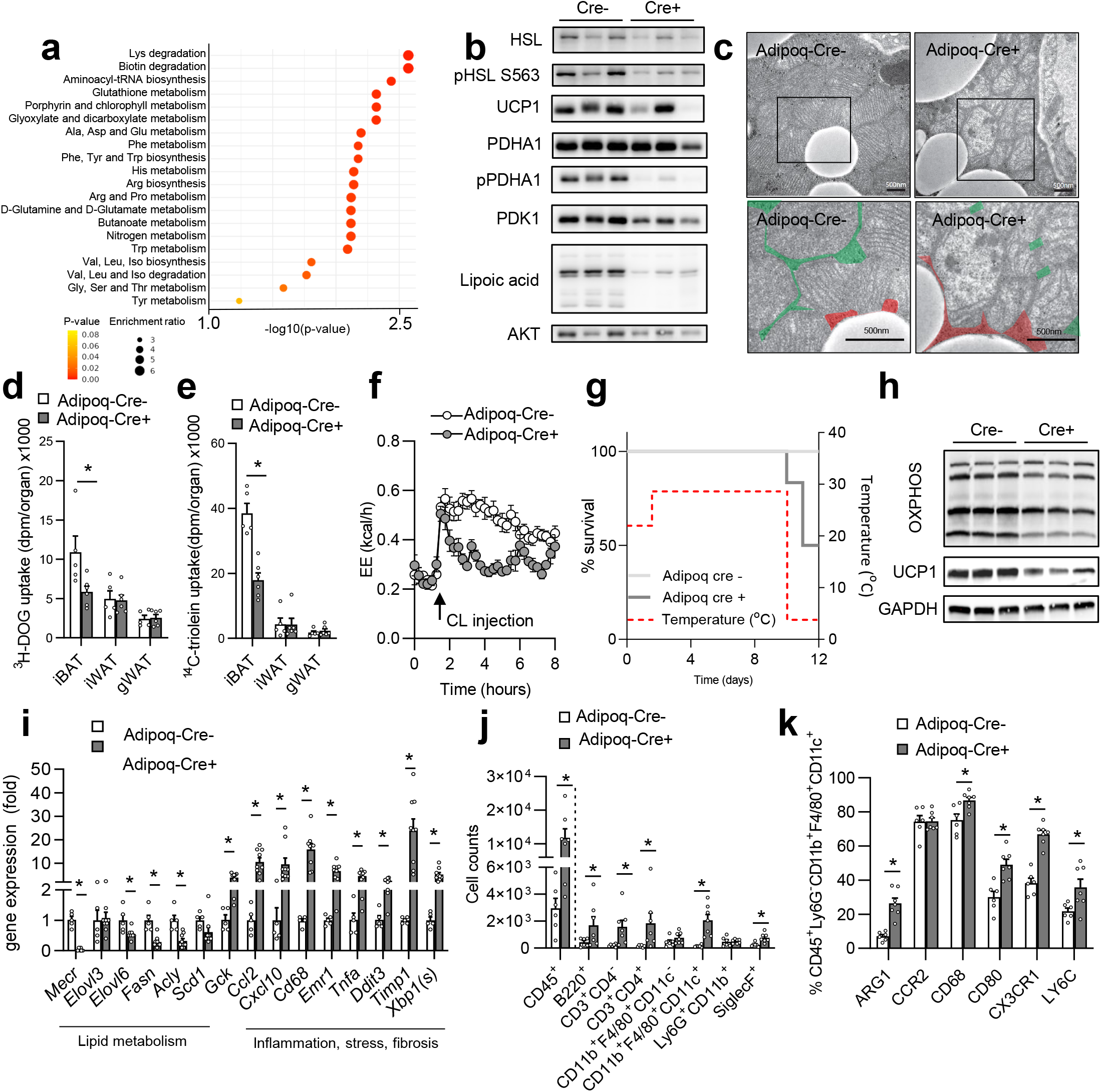
Loss of adipocyte Mecr leads to brown adipose tissue inflammation and dysfunctional thermogenesis. (a) Metabolic pathways enriched based on targeted metabolomics data of brown adipose tissue (n=6; for single metabolites see also Fig.S2b). (b) Western blot of brown adipose tissues (n=3). (c) Electron micrographs of brown adipose tissue (n=3) of *Mecr*^*flox/flox*^ *Adipoq-Cre* mice. (d) ^3^H-deoxyglucose (DOG) and (e) ^14^C-triolein tracer uptake into indicated organs after combined oral glucose and lipid gavage (n=5-6). (f) Energy expenditure of *Mecr*^*flox/flox*^ *Adipoq-Cre* mice injected with the beta3-adrenergic agonist CL316,243 at thermoneutrality (n=6). (g) Survival of *Mecr*^*flox/flox*^ *Adipoq-Cre* mice during cold exposure (experiment stopped at d3 of cold exposure). (h) Western blot of brown adipose tissues after 3d of cold exposure (n=3). (i) Gene expression of brown adipose at conventional housing temperature as well as (j) flow cytometry analysis of infiltrating cells and (k) phenotypic characterisation of BAT macrophages (n=6). Error bars indicate standard error of the mean (SEM). *= p<0.05 by Student’s T-Test.

### Liver-specific loss of *Mecr* leads to lipoylation depletion, impairment of PDH activity and early death

As *Mecr* is highly expressed in liver tissue (Fig.S1a) and the liver is a central organ for glucose and lipid handling, we generated *Mecr*^*flox/flox*^ *Alb-Cre* mice, to study the role of MECR for systemic lipid and glucose metabolism, Interestingly, the hepatocyte-specific deletion of *Mecr* led to globally affected mice with lower body weight (Fig.S4a), lower fasting blood glucose (Fig.S4b), cachexia-like adipose tissue wasting (Fig.S4c) as well as early death around the age of 14 weeks. For this reason, experimental interventions with the Cre^+^ homozygous floxed mice were performed before the 12^th^ week of age. For breeding, heterozygous floxed Cre^+^ mice were paired with homozygous floxed Cre^-^ mice because homozygous floxed Cre^+^ females did not survive until the end of pregnancy and homozygous floxed Cre^+^ males showed low fertility, likely due to failure to thrive. Interestingly, hepatocyte-specific *Mecr* deletion resulted in hepatomegaly, which was evident by an almost 2-fold increase of the percent liver-to-body weight ratio (Fig.S4c). Lipid analysis unveiled a rearranged hepatic lipidomic profile in the adult *Alb-Cre*^*+*^ mice (Fig.2a), with marked changes on triacylglycerol (TAG) (>70% reduction) and CE (>50% reduction), but also ceramides (CER) (25% reduction), SM (30% reduction) and to a lower extend all other investigated lipid classes (LPC, FFA, LPE, PC, PE ranging from 15%-25% reduction). As expected, in *Alb-Cre*^*+*^ mice with white adipose tissue wasting, white adipose tissue had lower TAG and higher phospholipids, most likely because the ratio of membrane-associated lipids to storage lipids is higher in their depots (Fig.3a). As observed by H&E staining, the livers of the *Alb-Cre*^*+*^ mice were characterized by enlarged hepatocytes in comparison to the wild type littermates (Fig.3b). Further, electron microscopy revealed that the hepatocytes were completely filled with swollen mitochondria (Fig.3c), features of early oncosis and cell death, which is described for acute liver failure (Bantel and Schulze-Osthoff, 2012). Following this observation, we next estimated the effect of MECR on mitochondrial function by seahorse-based oxygen consumption rate analysis in HepG2 cells treated with *MECR* siRNA, as primary hepatocytes of *Alb-Cre*^*+*^ mice were not viable. HepG2 cells treated with siRNA against *MECR* showed reduced oxygen consumption rates during basal respiration and uncoupled respiration already after 24h of siRNA treatment (Fig.3d), indicating a deleterious mitochondrial phenotype. Mitochondrial dysfunction is associated with cellular stress responses and indeed, upon hepatic deletion of *MECR*, gene expression analysis revealed high liver stress indicated by *Ddit3*, interestingly without markers of ER-stress (*Xbp1, sXbp1*). In line, we detected an increased expression of the pro-inflammatory cytokine *Tnfa* and the macrophage marker *Cd68* in the *Alb-Cre*^*+*^ mice, whereas other genes usually highly abundant in liver were hardly detectable (*Fasn, Ppara, Fgf21*) in adult mice (Fig.3e).

**Figure 3:**
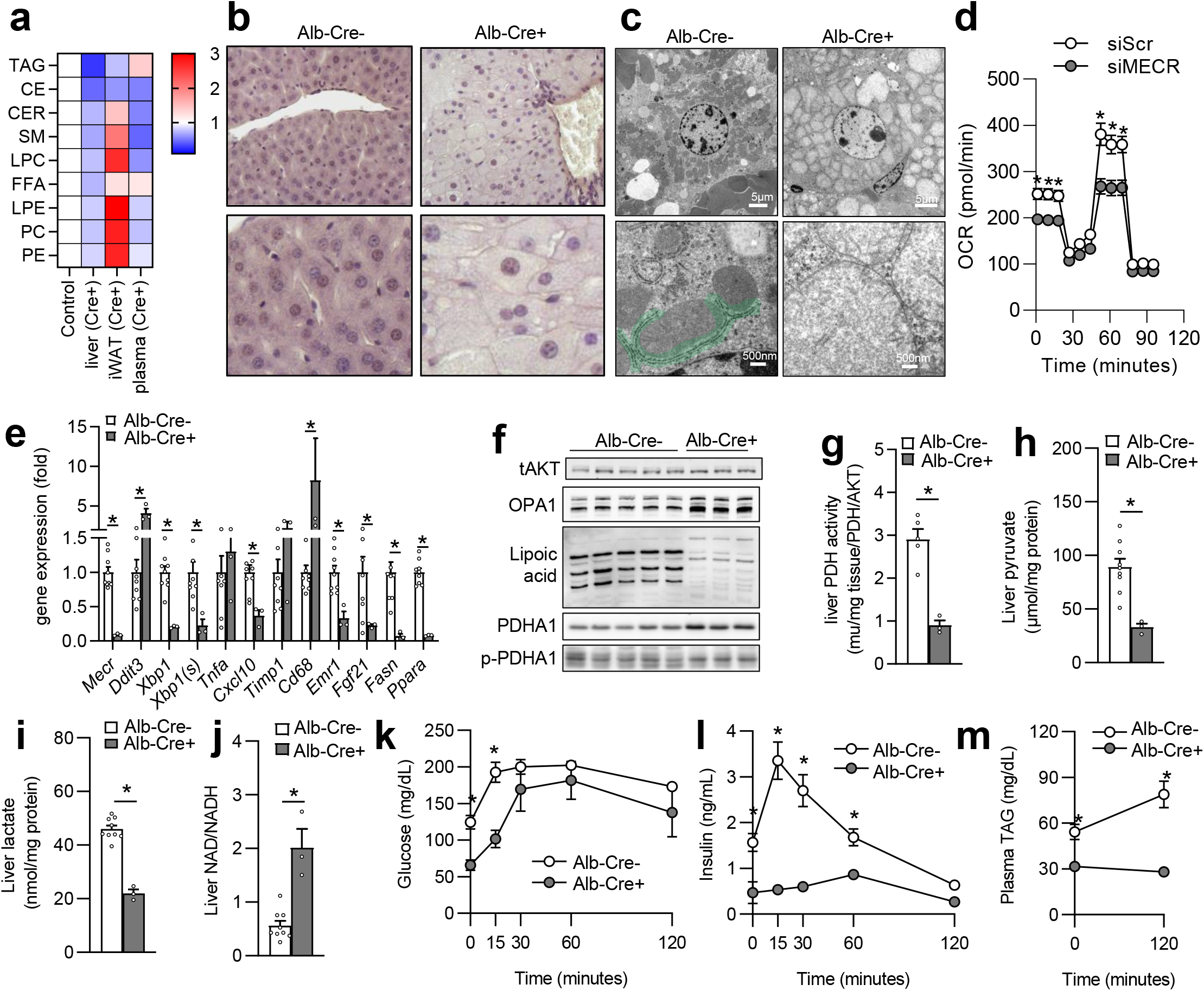
Liver-specific loss of Mecr leads to lipoylation depletion, impairment of PDH activity and early death. (a) LC/MS-based targeted lipidomics of liver, inguinal white adipose tissue (iWAT) or plasma from *Mecr*^*flox/flox*^ *Alb-Cre* mice (n=3-5).(b) H&E staining and (c) electron microscopy of livers from *Mecr*^*flox/flox*^ *Alb-Cre* mice (n=3). (d) Seahorse analysis of HepG2 cells transfected with either siMECR or control (n=12). (e) Hepatic gene expression analysis (n=3-9), (f) western blot of liver from *Mecr*^*flox/flox*^ *Alb-Cre* mice (n=3). (g) PDH activity (n=3-5), (h) liver pyruvate (n=3-9), (i) liver lactate (n=3-9), (j) NAD/NADH ratio of livers (n=3-9), (k) blood glucose (n=3-9), (l) plasma insulin (n=3-9), (m) plasma triglycerides and ^14^C-deoxyglucose uptake from *Mecr*^*flox/flox*^ *Alb-Cre* mice gavaged with a combined radioactive glucose and lipid load (n=3-9). Error bars indicate standard error of the mean (SEM). *= p<0.05 by Student’s T-Test.

Western blot analysis of the livers showed that, despite the massive enlargement of the mitochondria, oxidative phosphorylation complex levels were only mildly affected on protein level (Fig.S2e). However, markers of intracellular energy availability such as lysine-malonylation and –succinylation were dysregulated on protein level with reductions in *Alb-Cre*^*+*^ mice (Fig.S4d). Of note, the reduced phosphorylation of AKT in the *Alb-Cre*^*+*^ mice, pointed towards an attenuated insulin signaling. Importantly, as indicated by the impaired protein lipoylation pattern in the liver of *Alb-Cre*^*+*^ mice (Fig.3f), loss of liver MECR most likely decreased lipoic acid synthesis. Additionally, and in line with the formation of megamitochondria, the cleaved form of OPA1, an important protein that regulates mitochondrial fusion/fission dynamics, was particularly elevated in the livers of *Alb-Cre*^*+*^ (Fig.3f).

Since lipoic acid is a crucial cofactor for PDH, the levels of this enzyme were further investigated. Despite elevated hepatic protein levels of the PDHA1 in the *Alb-Cre*^*+*^ mice (Fig.3f), the PDH activity was significantly diminished (Fig.3g), most likely due to almost absent protein lipoylation (Fig.3f). Since the phosphorylated form of PDHA1 was not significantly altered (Fig.3f), the elevated protein levels of PDHA1 might stem from a feedback mechanism to counteract its attenuated activity. Despite the lower PDH activity, hepatic pyruvate did not accumulate in *Alb-Cre*^*+*^ mice, but was rather decreased (Fig.3h), which might in part be explained by the hypoglycemia of the mice, which generally leads to lower glycolysis (Unger, 1985). Besides its degradation by PDH, pyruvate can alternatively be converted into lactate by lactate dehydrogenase, which regenerates NAD^+^ for ongoing glycolysis. Hepatic lactate levels were also lower in *Alb-Cre*^*+*^ mice (Fig.2i) overall suggesting lower glycolytic activity. We next speculated, that glycolysis might be reduced due to a lack of NAD^+^ which cannot be regenerated from pyruvate. Nevertheless, the hepatic ratio of NAD^+^ to NADH was critically increased (Fig.3j) in *Alb-Cre*^*+*^ mice, arguing that enough NAD^+^ would be available for ongoing glycolysis and that NAD^+^ levels are not the reason for the failure of glycolytic activity. Of note, also NADP^+^/NADPH was increased and therefore in principle the pentose phosphate pathway was impaired as well (Fig.S4e), whereas ATP levels were similar in both genotypes (Fig.S4f). In addition, enzyme activity of alpha-ketoglutarate dehydrogenase, which like PDH also relies on lipoic acid as cofactor, was also significantly reduced in *Alb*-Cre^+^ mice (Fig.S4g).

To characterize the postprandial metabolic capacity, we performed a combined oral glucose and fat tolerance test. Interestingly, *Alb-Cre*^*+*^ mice showed not only a lower basal glucose level, but also reduced levels during the oral glucose tolerance test (Fig.3k). Surprisingly, the insulin levels of the *Alb-Cre*^*+*^ mice were almost unchanged during the glucose load (Fig.3l), indicating a lower insulin dependency of glucose clearance or an increased insulin sensitivity. Plasma triglyceride levels were elevated during lipid load in *Alb-* Cre^-^, but not in *Alb-*Cre^+^ mice, which also presented lower basal triglyceride (Fig.3m) and cholesterol (Fig.S4h) levels.

Overall, our data suggest that the reduced NADH/NADPH formation capacity and reduced dehydrogenase activities observed in *Alb-Cre*^+^ mice might be important contributors to fatal cause of liver dysfunction ultimately resulting in disturbances in systemic glucose and lipid metabolism.

### Functional analysis of human *MECR* gene variants reveal disturbed glucose and lipid homeostasis

A male patient in his 20s presented clinically with childhood onset optic nerve atrophy and gait ataxia, dystonia, generalized myoclonia and choreoathetoid movements. Trio-whole exome sequencing revealed a biallelic mutation in *MECR*. It uncovered a loss-of-function mutation c.553delG (p.D185I*fs*\*25), and a c.772C>T (p.Arg258Trp) previously published pathogenic missense variant (Heimer et al., 2016) in compound-heterozygous state. To characterize the impact of these patient variants on cellular function, we overexpressed wild-type *MECR*, as well as both variants in HEK cells. First, we assessed PDH activity as PDH is known to depend on lipoic acid as a cofactor. Cells transfected with wild-type *MECR*, but not with one of the two patient variants, showed increased PDH activity (Fig.4a). Next, we assessed glucose uptake upon overexpression of wild-type and variant *MECR*. Of note, overexpression of both variants, but not the wild-type transcript, led to higher uptake of ^3^H-deoxyglucose into the cells (Fig.4b), suggesting a dominant negative effect of each *MECR* variant on mitochondrial catabolism and a subsequent higher need for anaerobic glycolysis to compensate. To assess the functional impact of mtFAS on cellular lipid metabolism, we performed LC/MS-based targeted lipidomic analysis of HEK293T cells transfected with wild-type *MECR* and the patient *MECR* variants. Interestingly, we found that membrane lipids, such as (lyso)-phosphatidylcholine ((L)PC), (lyso-) phosphatidylethanolamine ((L)PE), and sphingomyelin (SM), were lower upon overexpression of the patients *MECR* variants (Fig.4c; Fig.S5a). Overexpression of wild type *MECR* caused higher abundance of cholesterylester (CE) species containing palmitate and oleate residues (Fig.4d), which reflect fatty acid species described to be specifically derived from cytosolic *de novo* lipogenesis (Heeren and Scheja, 2021; Worthmann et al., 2022). While the frameshift variant p.D185I*fs*\*25 showed similar lipid species concentrations compared to cells transfected with the empty vector control, the missense variant p.Arg258Trp showed a tendency towards higher levels of indicated CE species as well, indicating a potential residual activity of the MECR^R258W^ variant. Evaluation of the mouse models revealed that cholesterylester species in plasma were not influenced by adipose tissue mtFAS deficiency (Fig.4e), but liver (Fig.4f). However, the lipid class itself might not be specific to CE, as e.g., in BAT lower carbon fatty acid residues in PC were decreased, whereas longer and higher desaturated fatty acids were increased (Fig.S5b). In line, the patient with biallelic *MECR* variants showed reduced levels of palmitate and palmitoleate residues in TAGs and CEs compared to plasma of healthy sex- and age-matched controls (Fig.S5c), which might be partially influenced by the low mtFAS level. Based on these observations, we hypothesized that mtFAS and subsequently identified mtFAS-associated lipids might also be disturbed in patients with confirmed other mitochondriopathies.

**Figure 4:**
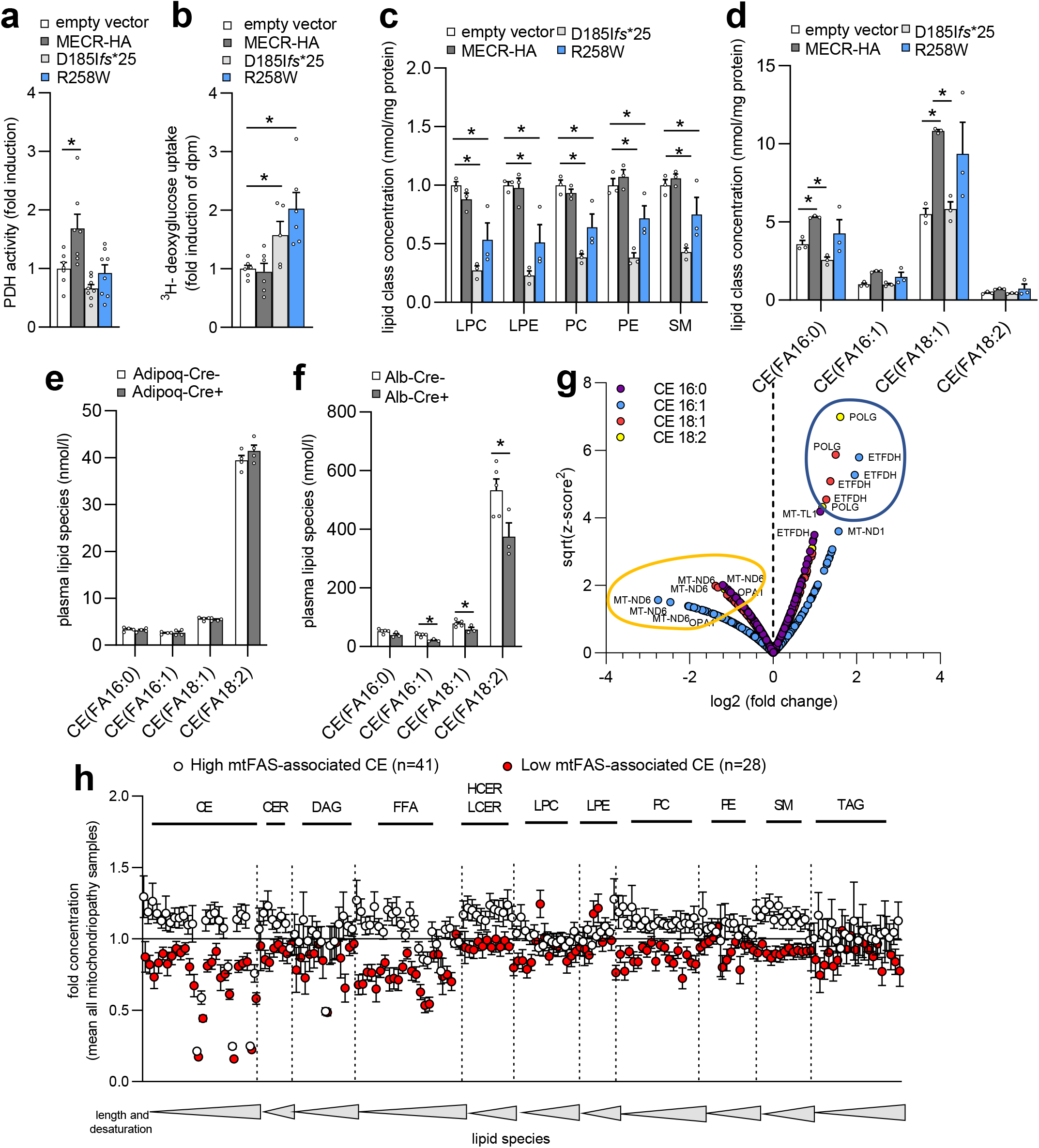
Functional characterization of variants in MECR reveal disturbed glucose and lipid homeostasis. HEK293T cells overexpressed either *MECR* wild type or indicated variants for 24h and (a) PDH activity (n=7-8), (b) ^3^H-deoxyglucose uptake (n=5-6) or (c-d) targeted lipidomics (n=3) were performed. (e) Plasma lipidomics of *Adipoq-Cre* or (f) *Alb-Cre* mice. (g) Lipidomic analysis of 300 plasma samples from patients with genetically confirmed mitochondriopathies were analyzed with regard to MECR overexpression-associated lipid species found in (d) and were grouped based on the genes *MT-ND6 (low), ETFDH* and *POLG* (high) into low and high mtFAS-associated CE patients, which were grouped in (f) with regard to their plasma lipid classes and the respective length and desaturation. Error bars indicate standard error of the mean (SEM). *= p<0.05 by two-way ANOVA and Fishers-LSD Test.

Therefore, we screened a cohort of over 2000 patients with confirmed or suspected mitochondrial disease from the mitoNET consortium and selected adult cases, who had a genetically confirmed diagnosis and took part in biobanking (n=300). We performed lipidomic analysis of their plasma samples focusing on the respective mtFAS-associated CEs identified to be increased by *MECR* expression. We found that compared to the cohorts mean CE abundance especially patients with pathogenic variants in *MT-ND6* (n=28) showed lower mtFAS-associated CEs whereas patients with pathogenic variants in *POLG* and *ETFDH* showed higher mtFAS-associated CEs (n=41) (Fig.4g). Based on the abundance of these CEs we defined subgroups with low and high mtFAS-associated CE amongst these patients, which of note, are also influenced by many other factors. Next, plotting the plasma concentration of all lipid species analyzed (sorted by their length and degree of desaturation) in the low and high mtFAS subgroup revealed that especially CEs, free fatty acids (FFA), hexosyl ceramides (HCER), lactosyl ceramides (LCER), PC and SM were distinguishing these two groups (Fig.4h). In sum, this indicates that multiple lipids are affected by the *MECR*, that specific CE lipid species may be affected in vivo by mtFAS and thus it is tempting to speculate that also other, currently non-mtFAS-associated mitochondriopathies may show an intrinsic dysregulation in mtFAS activity.

## Discussion

Our study shows that mtFAS reflects an important pathway for systemic lipid and glucose metabolism by regulating the energy transformation in liver and adipose tissues. As critically involved in lipoic acid production, mtFAS reflects an important pathway to fully break down and transform glucose into reduction equivalents or precursors for other metabolic pathways such as fatty acid synthesis. In adipose tissue, loss of *MECR* leads to reduced membrane lipids, cigar-shaped mitochondria, reduced PDH activity, increased lipolysis and higher VLDL production. In a model of diet-induced obesity, adipocyte specific deletion of *Mecr* reduced glucose tolerance, insulin sensitivity and nutrient uptake into WAT. Moreover, in brown adipose tissue *Mecr* deficiency caused parenchymal failure as indicated by reduced beta adrenergic response, cold intolerance, and severe BAT inflammation with induction of profibrotic solution markers. We found that loss of *Mecr* in the liver caused severe hepatic failure with decreased hepatic storage and membrane lipid content, enlarged megamitochondria, swollen hepatocytes, deranged transcription, reduced PDH activity, and lower production of reduction equivalents ultimately resulting in hypoglycemia, hypolipoproteinemia, and disturbed glucose uptake. These data suggest that appropriate mtFAS substantially contributes to energy homeostasis and organ function.

Mechanistically, our study revealed that loss of *MECR* resulted in the loss of protein lipoylation within the respective organ. As lipoic acid is one of the key cofactors for PDH activity, subsequently its activity was reduced. As lipoic acid cannot be transported into mitochondria because they lack respective transporters (Tajima et al., 2019), lipoic acid supplementation is likely to be no promising therapeutic option. However, for some individuals with biallelic *MECR* variants benefits have been observed (Liu et al., 2021). Another yet to be addressed mechanism of mtFAS for mitochondrial function and its subsequent effects on systemic energy metabolism might be its importance for the assembly of respiratory complexes. In particular, *Mecr* derived acyl-chains have been described to facilitate the assembly of respiratory complex I independent from lipoic acid (Tanvir Rahman et al., 2023). The broader relevance of our findings implicates that mtFAS might be affected in physiologic and pathophysiologic states of liver and adipose tissues. We show that mtFAS regulates the uptake and transformation of nutrient derived energy into reduction equivalents such as NADH and NADPH.

These may be used to fuel fatty acid synthesis directly or alternatively, fatty acid synthesis might be regulated by mtFAS via the mitochondrial production of citrate/acetyl-CoA precursors for *de novo* lipogenesis (Heeren and Scheja, 2021; Worthmann et al., 2022).

mtFAS seems to be a key pathway needed for functional nutrient metabolism in the respective organ with life threatening loss of function in mitochondria-rich organs, such as BAT and liver. In contrast, humans with biallelic mutations in *MECR* do not show liver failure, whereas mice with tissue specific loss do. In our patient, we speculate that a residual activity of the missense variant might protect from early death and so far, no biallelic loss-of-function patient has been described (Heimer et al., 1993). However, it might also be possible that components of human food, which are not available in standard rodent diet might be able to partly compensate for the loss of mtFAS-derived products.

We showed *in vitro* that mtFAS seems to be important for the abundance of long-chain fatty acids esterified to cholesterol. This was also phenocopied in plasma of *Alb-Cre*^*+*^ but not in *Adipoq-Cre*^*+*^ mice, indicating a direct secretion of the respective lipid species from the liver. However, in BAT, PC lipidomic signature showed distinct changes towards longer fatty acid residues and higher desaturation and in a patient with biallelic MECR variants, specific TAG species with similar fatty acid residues were decreased indicating that residues of complex lipids are influenced and that the changes are not limited to a specific complex lipid class and might vary by cell type and/or organ. Whether these fatty acid residues are produced by mtFAS itself or whether mtFAS facilitates cytosolic *de novo* lipogenesis or exogenous lipids is unclear yet. Interestingly, CE containing the mtFAS-associated fatty acid residues analyzed in HEK cells were also identified to be lower in patients with certain mitochondriopathies, caused by variants in *MT-ND6*, and increased in mitochondriopathy patients caused by variants in *ETFDH* and *POLG*. It will be of interest to evaluate whether mtFAS is dysregulated in these mitochondriopathies subsequently altering levels of these lipids or whether also acquired mitochondrial dysfunction described e.g., in patients with diabetes (Pinti et al., 2019) or chronic liver disease (Mansouri et al., 2018) might affect mtFAS and *vice versa*.

In conclusion, we describe mtFAS to be essential for mitochondrial and metabolic health largely affecting liver and adipose biology. This study gives insights that by setting the metabolic transformation capacity, mtFAS reflects one of the major pathways needed for whole body glucose and lipid tolerance.

## Data Availability

All data produced in the present study are available upon reasonable request to the authors.

## Acknowledgements

We are thankful to Laura Ehlen, Katrin Rading, Verena Rickassel and Meike Kröger for excellent technical support. We also thank Julia Scarpa and Silke Becker for support at the Metabolomics and Proteomics Core of Helmholtz Zentrum München. We thank Kristl Claeys, Markus Schülke-Gerstenfeld, Cornelia Kornblum, Jochen Schäfer, Leila Scholle, Christian Knop, Daniella Karall and Ludger Schöls for participation in mitoNET biosampling of the used samples. We also thank Julia Scarpa and Silke Becker for support at the Metabolomics and Proteomics Core of Helmholtz Zentrum Munich. C.S. was supported by the Werner-Otto-foundation, by the DFG (SCHL2276/2-1), the Mühlbauer foundation and by the University Medical Center Hamburg Eppendorf medical faculty (TDM-21/06). A.W. is supported by the University Medical Center Hamburg Eppendorf medical faculty (TDM-21/06; NWF-20/07) and the Mühlbauer-foundation. F.H. is supported by the Werner-Otto-foundation. J.H. (P05), K.A.D. (S02), C.S. (P15), L.B. (P07) and A.W. (P07) are supported by a grant funded by the DFG (450149205-TRR333/1). A.B. is supported by the Deutsche Forschungsgemeinschaft Sonderforschungsbereich 1123 (B10), the Deutsches Zentrum für Herz-Kreislauf-Forschung Junior Research Group Grant, the European Research Council Starting Grant “PROTEOFIT”, and the DFG Priority Programme on ferroptosis SPP2306. In addition, A.W., L.B. and J.H. are supported by the State of Hamburg (LFF-FV75).

## Author contributions

F.H., I.E. and A.W. generated data, analyzed data and wrote the manuscript. S.K., A.T.B., M.H., K.D., I.L., L.B., C.P., J.H., A.B. and C.K. generated or analyzed data and edited the manuscript. B.B., C.N., T.K. and H.P. analyzed and supervised the mitoNET consortium and biobank and edited the manuscript. C.S. initiated and designed studies, analyzed data, directed overall execution of studies and wrote the manuscript. C.S is the guarantor of this work and, as such, had full access to all the data of the study and takes responsibility for the integrity of the data and the accuracy of the data analysis.

## Methods

### Study participants

Trio-whole-exome sequencing on lymphocytes was performed in the index patient. For lipidome analysis, plasma of 300 patients with genetically confirmed mitochondrial disease was used from the mitoNET cohort (www.mitoNET.org), which is a multicentric German network for mitochondrial diseases including a biobank for mitochondrial disease. All analyses were carried out with written informed consent and the study was approved by the local medical ethics committees.

### Animal models

All animal studies were performed with permission of the Animal Welfare Officers at University Medical Center Hamburg-Eppendorf. B6(Cg)-Mecr^tm1c(EUCOMM)Wtsi/WtsiCnbcOulu^ mice were purchased from the European Mouse Mutant Archive (EMMA) provided by A. Kastaniotis (University of Oulu) and were crossed to Alb-Cre or Adipoq-Cre lines purchased from Jackson Laboratories. Mice on a high fat diet were fed a 60% kcal from fat diet (EF Bio-Serv F3282, Ssniff®). Age and weight-matched male mice (8-20 weeks) were used. Routinely, mice were kept in single cages during adaptation to indicated environmental temperatures in Memmert climate chambers with ad libitum access to food and water. Blood for EDTA plasma measurements was collected by tail vein blood withdrawal or cardiac puncture of anaesthetized mice.

### Indirect calorimetry

Indirect calorimetry was performed using the PROMETHION systems (Sable Systems International) in a temperature- and humidity-controlled chamber. During the experiment, all mice were housed in single cages under a 12 h light: 12 h dark cycle and had ad libitum access to food and water. *Mecr*^*flox/flox*^*Adipoq-Cre*^*-*^ and *Mecr*^*flox/flox*^*Adipoq-Cre*^*+*^ as well as *Mecr*^*flox/flox*^*Ucp1-Cre*^*-*^ and *Mecr*^*flox/flox*^*Ucp1-Cre*^*+*^ mice were kept initially for 2 days at the system under conventional housing temperature (22°C) and then kept at thermoneutral (30°C, 7 days) followed by cold (4°C, 3 days) ambient temperatures for the indicated time period. On the last day of thermoneutral housing, mice were injected intraperitoneally with 100uL of the selective beta3-adrenegic receptor agonist CL316,243 (Cayman Chemicals) at a concentration of 0.2 mg/mL in 0.9% saline. Oxygen consumption and carbon dioxide production were measured every 15 min.

### Cell culture

For lipid analysis, HEK293T cells were seeded into each well of 6-well plates and incubated at 37°C for 24h with plating medium (DMEM, 10% FCS, 1% Anti-Anti). Cells were then transfected using JetOptimus DNA transfection reagent (Polyplus). Per 6-well a mix of 2 μg Plasmid DNA (pcDNA 3.1(+)-N-HA; MECR pcDNA 3.1; MECR_OHu_c.772CT; MECR_OHu_c.553delG), 200 μl buffer and 3 μl JetOptimus reagent was added. Media was changed to fresh plating medium after 5h of incubation at 37°C. 24h after transfection cells were washed with ice cold PBS containing free fatty acid-free BSA (33.3 mg/ml; pH 7). After an additional washing with ice cold PBS, cells were harvested in 1 ml PBS using a cell scraper and transferred into 2 ml Eppendorf reaction tube. Cell suspensions were spun down (650 rcf; 5 min; 4°C), supernatant discarded, and pellets stored at -80°C ready for lipid analysis.

### Extracellular flux analysis (Seahorse)

Oxygen consumption rate was analyzed using a Seahorse XFe24 Analyzer (Agilent). 40,000 HepG2 cells per well were transfected with either SMARTPool ON-TARGETplus Human MECR siRNA (Dharmacon) or scramble control using Lipofectamine™ RNAiMAX (ThermoFisher Scientific) according to the manufacturer’s instructions. 24h after transfection, a Mito Stress Test using 1μM oligomycin, 0.5μM FCCP (Carbonyl cyanide-p-trifluoromethoxyphenylhydrazone) and 0.5μM rotenone/antimycin-A (all Sigma-Aldrich) was performed. The measurements were performed according to the Mito Stress user guide provided by Agilent.

### Gene expression

RNA was isolated from tissues using 1 ml peqGEOLD TriFast (Peqlab) per sample. Tissues were homogenized with Tissue Lyser (Qiagen), 200 μl chloroform were added and a phase separation was performed (centrifugation 15 min; 13,000 rpm; 4°C). RNA containing aqueous phase was mixed with 70 % ethanol (1:1) and RNA was purified by NucleoSpin RNA II Kit (Machery-Nagel). After RNA isolation, cDNA was transcribed with High-Capacity cDNA Archive Kit (Applied Biosystems). RNA samples concentration and quality was measured with absorbance plate reader (EPOCH; BioTek). Gene expression was assessed using Real Time PCR as SYBR green (Applied Biosystems). Data were normalized to the gene expression levels of the housekeeper *Tbp* for mice or *TAF1* for human samples. Samples without detectable housekeeping or target gene amplification were excluded. SYBR green assays (supplemental table) were designed to avoid single nucleotide polymorphism sites and PCR products were designed to span exons, which preferably have long introns to reduce unspecific amplification of possible genomic DNA contamination.

### Western blotting

Perfused organs were harvested and homogenized with a TissueLyzer (Qiagen) at 20 Hz for 2x 3 min in 10x (v/w) RIPA-buffer (50 mM Tris-HCl pH 8; 150 mM NaCl; 5 mM EDTA; 1% NP-40; 0.5 % Na-Deoxycholate; 0.1 % SDS) supplemented with cOmplete Mini Protease Inhibitor Cocktail Tablets (Roche), PhosStop Tablets (Roche) and HDAC-inhibitor (Santa Cruz). Lysates were clarified by centrifugation (14,000 rpm, 15 min, 4°C) and protein concentrations of supernatant were measured with BCA method (Thermo Scientific). For white adipose tissues supernatant was taken using a syringe and a 27G needle to avoid contamination from the fat layer. Supernatants were then supplemented with 4x laemmli buffer (33 % glycerol; 0.3 M DTT; 6.7 % SDS; 0.01 % bromophenol blue; 80 mM Tris-HCl pH 6.8) and proteins were separated with SDS-PAGE electrophoresis. Bio-Rad 4-20% gradient gels were loaded with 20 μg protein per well and run at 150 V and immediately transferred to a 0.2 μm PVDF membrane using Bio-Rad Transblot Turbo Transfer System. After blocking the membranes for 1h at room temperatures using 5 % non-fat dry milk in TBS-T buffer (20mM Tris-HCL, pH 7.4, 150 mM NaCl, 0.1 % Tween-20), membranes were washed thrice for 5min in TBS-T buffer. Following membranes were incubated overnight at 4 °C in primary antibody solution (5 % BSA in TBS-T buffer) containing the appropriate antibodies. Membranes were washed as previously and incubated for 1h at room temperature with goat anti-rabbit or horse anti-mouse IgG horseradish peroxidase (HRP) secondary antibody (cell signalling; 1:2500 in 5% non-fat dry milk in TBS-T). After final washing signals were detected using ECL reagent (WesternBright; Advansta) with a 2 min incubation, after which the membranes were imaged using ChemiDoc MP Imaging System (Bio-Rad laboratories).

| Primary antibodies | Company | Cat# |
| --- | --- | --- |
| Rabbit polyclonal anti-AKT | Cell Signalling | 9272 |
| Rabbit polyclonal anti Phospho-AKT (Ser473) | Cell Signalling | 9271 |
| Rabbit monoclonal anti Phospho-AKT(Thr308) | Cell Signalling | 13038 |
| Mouse OXPHOS WB cocktail | Abcam | ab110413 |
| Rabbit monoclonal anti-OPA1 | Cell Signalling | 80471 |
| Rabbit polyclonal anti-lipoic acid | Abcam | ab58724 |
| Rabbit monoclonal anti-PDHa1 | Cell Signalling | 3205 |
| Rabbit monoclonal anti-Phospho-PDHa1 | Cell Signalling | 37115 |
| Rabbit polyclonal anti-UCP1 | abcam | ab10983 |
| Rabbit monoclonal anti-Malonyl-Lysine | Cell Signalling | 14942 |
| Rabbit polyclonal anti-Succinyl-lysine | PTM Biolab | PTM-401 |
| Mouse monoclonal anti-GAPDH | Abcam | ab8245 |
| Rabbit polyclonal anti-Phospho-HSL (Ser563) | Cell Signalling | 4139 |
| Rabbit polyclonal anti-HSL | Cell Signalling | 4107 |
| Rabbit monoclonal anti-gamma Tubulin | Abcam | ab179503 |
| Rabbit polyclonal anti-MECR | Proteintech Europe | 14932-1-AP |

### Lipidomic analysis

Plasma lipidomic analysis was performed using the Lipidyzer™ Platform from SCIEX. Briefly, plasma and cell samples were spiked with Lipidyzer™ Internal Standards (SCIEX) and lipid extraction was performed employing an adjusted MTBE/methanol extraction protocol. Extracted lipids were concentrated and reconstituted in a mixture of dichloromethane (50): methanol (50) containing 10mM ammonium actetate. Separation and targeted profiling of lipid species was performed combining differential mobility spectrometry and a QTRAP® system (QTRAP® 5500; SCIEX). Quantification of lipids was conducted by the Lipidyzer™ software (Lipidomics Workflow Manager software; SCIEX) employing specific multiple reaction monitoring transitions.

### Targeted metabolomics

Targeted metabolomics of adipose tissue extracts was performed using the Absolute*IDQ*™ p180 Kit (BIOCRATES Life Sciences AG, Innsbruck, Austria), as previously described (Artati et al., 2022). This method combines liquid chromatography-electrospray ionization-tandem mass spectrometry (LC-ESI-MS/MS) and flow injection-electrospray ionization-tandem mass spectrometry (FIA-ESI-MS/MS) to quantify 188 metabolites, including free carnitine, 39 acylcarnitines, 21 amino acids, 21 biogenic amines, sum of hexoses, 90 glycerophospholipids, and 15 sphingolipids. Amino acids and biogenic amines are quantified in the LC run, based on scheduled multiple reaction monitoring measurements (sMRM). All other metabolites are quantified in the FIA run, based on MRM analyses. The LODs were set to three times the values of the zero samples (EtOH/PB 80/20 v/v). The assay procedures of the Absolute*IDQ*™ p180 Kit and the tissue preparation and have been described in detail previously (Zukunft et al., 2018). In brief, tissue homogenates were always prepared freshly as follows: Frozen murine white or brown adipose tissue samples were weighted into homogenization tubes with ceramic beads (1.4 mm). For metabolite extraction, <u>to each 1 mg</u> of frozen murine adipose <u>tissue</u> were added <u>12 μL</u> of a to 4 °C cooled mixture of <u>ethanol/phosphate buffer</u> (80/20 v/v). Tissue samples were homogenized using a Precellys24 homogenizer (PEQLAB Biotechnology GmbH, Germany) 4 x for 20 s at 5,500 rpm and 5-10 °C, with 30 s pause intervals to ensure constant temperature, followed by centrifugation at 10,000 x g for 5 min. Subsequently, 10 μL of the supernatants were analyzed with the p180 assay. Data evaluation for quantification of metabolite concentrations and quality assessment were performed with the software MultiQuant 3.0.1 (SCIEX) and the Met*IDQ*™ software package, which is an integral part of the Absolute*IDQ*™ Kit. Metabolite concentrations were calculated using internal standards and reported either in pmol/mg for wet tissue or μmol/L (μM) for tissue homogenate. In addition to the investigated samples, five aliquots of a pooled reference plasma (Ref_Plasma-Hum_PK3) were analyzed on each kit plate. The results of these reference plasma aliquots can be used for calculation of potential batch effects and data normalization (of different studies). The long-time stability of plasma metabolites during storage at -80 °C and the performance of our targeted-metabolomics platform have been evaluated in (Haid et al., 2018).

### PDH activity assay

Pyruvate dehydrogenase activity was measured with PDH activity assay kit (Sigma Aldrich). Liver tissues were homogenized in 10 x PDH assay buffer (v/w) with tissue lyzer (Qiagen) and have been clarified via centrifugation. 15μl of lysate have been used for measurement. White adipose tissues were homogenized in 5-times the amount of PDH assay buffer (5:1 v/w), then clarified and 30μl of lysates have been used for measurement. Following measurement has exactly been performed as described in the kit’s manual. Generated amount of NADH was measured at 450 nm with a photometer (Synergy H1 BioTek) every 5-10 min and PDH activity was calculated as one unit of pyruvate dehydrogenase is reported as the amount of enzyme that will generate 1.0 μmole of NADH per minute at pH 7.5 at 37 °C.

### Metabolic tracer study

For oral glucose and fat tolerance tests (OGFT) was performed as described earlier (Schlein et al., 2016). Briefly, mice were fasted for 4 hours before receiving an oral gavage of 200 μl of a glucose-lipid-emulsion containing glucose (2 g/kg body weight) and triacylglycerol-rich lipoproteins (80 mg triacylglycerol/kg body weight) traced with 3H-deoxyglucose (0.72 MBq/kg body weight) and 14C-triolein (0.15 MBq/kg body weight). At indicated timepoints blood for EDTA plasma measurements was collected by tail vein withdrawal and Glucose was measured in tail blood using Aviva AccuCheck glucose sticks (Roche). After 2 hours, mice were anaesthetized (xylazine/ketamine), blood collected by cardiac puncture and then systemic perfused with PBS-heparin (10 U/ml) via left heart ventricle. Organs were harvested and dissolved in 10x (v/w) Solvable (Perkin Elmer), and radioactivity (in dpm) was measured by scintillation counting using a Perkin Elmer Tricarb Scintillation Counter.

### Flow cytometry analysis

For the analysis of the infiltrating cells in the adipose tissue as well as the characterization of the macrophages, the mice were cardiac-punctured and perfused with PBS. Gonadal white as well as interscapular brown adipose tissues were collected and kept in PBS on ice for further processing. Afterwards adipose tissue was minced into small pieces on ice and incubated for 35 min at 37°C while shaking in 10 mL (WAT) or 8 mL (BAT) digestion buffer (HBSS, 1mg/mL Collagenase I (Worthington), 0.5% FCS, 1% 0.2M MgCl2, 0.4% 0.5M CaCl2). EDTA (final concentration 10 mM) was supplemented to the suspension and incubated for additional 10 min. Subsequently, cells were filtered over a 100 μm filter and centrifuged at 500 x g 10 min 4°C. The floating fraction was discarded. The pelleted cells were incubated in 1 mL red blood cell lysis buffer for 7 min at room temperature before stained for flow cytometry analysis. For this, cells were in incubated with Fc-block (BioLegend, anti-CD16/CD32) for 15 min at 4°C. Afterwards, cells were washed with PBS/2 % FCS and incubated with a surface-epitope antibody cocktail for 35 min at 4°C in the dark. For the analysis of intracellular epitopes, cells were fixed and permeabilized using the Foxp3/Transcription Factor staining Buffer Set (eBiosciences) and incubated for 35°minat 4°C in the dark with the intracellular antibody cocktail diluted in permeabilization buffer. Fluorescence of the stained cells were measured at the LSRII (BD Biosciense) and data were analyzed using the Flowjo software (Tree Star).

### Triacylglycerol, cholesterol and free fatty acid measurements

For plasma triglyceride and cholesterol analysis multi-purpose kits for cholesterol FS and triglycerides FS (Diasys Diagnostics) or free fatty acids (WAKO) were utilized using EDTA plasma or cell culture supernatant.

### Histochemistry

Hematoxylin-Eosin staining was performed on paraffin-embedded tissues using standard procedures. Briefly, sections (5 mm) were cut on a Leica Microtome and mounted on Histobond slides (Marienfeld-Superior). After de-paraffinization and rehydration, slides were incubated for 2 min in Hem-atoxilin, then blued under running tap water for 10 min, and counterstained with 0.5% Eosinfor 15 s. Afterward, the slides were de-hydrated and mounting was performed using Eukitt. Images were taken using Zeiss ApoTome.

### Electron microscopy

For electron microscopy mice were sacrificed with a lethal dose of ketamin/xylazine injection anesthesia and perfused with PBS containing 10U/ml heparin. Organs were cut and directly transferred into fixative (4% PFA, 1% GA in PBS) and stored at 4°C. Then, tissues were dissected with a razor blade and rinsed three times in 0.1 M sodium cacodylate buffer (pH 7.2–7.4) and osmicated using 1% osmium tetroxide in cacodylate buffer. Following osmication, the samples were dehydrated using ascending ethyl alcohol concentration steps, followed by two rinses in propylene oxide. Infiltration of the embedding medium was performed by immersing the pieces in a 1:1 mixture of propylene oxide and Epon and finally in neat Epon and hardened at 60 °C. Semithin sections (0.5 μm) were prepared for light microscopy mounted on glass slides and stained for 1 minute with 1% Toluidine blue.

Ultrathin sections (60 nm) were cut and mounted on copper grids. Sections were stained using uranyl acetate and lead citrate. Thin sections were examined and photographed using an EM902 (Zeiss) electron microscope.

### Statistical methods

Data are expressed as mean ± S.E.M. Comparisons of two groups were examined using Students T-Test or using Mann-Whitney test (flow cytometry data).Comparison of three or more groups were analyzed using ANOVA. GraphPad Prism and Microsoft Excel were used for all statistical analyses. The statistical parameters can be found in the figure legends. P < 0.05 was considered to be statistically significant.

