## Supplemental Figures for "Mitochondrial fatty acid synthesis is essential for coordinated energy transformation"

### Fig.S1

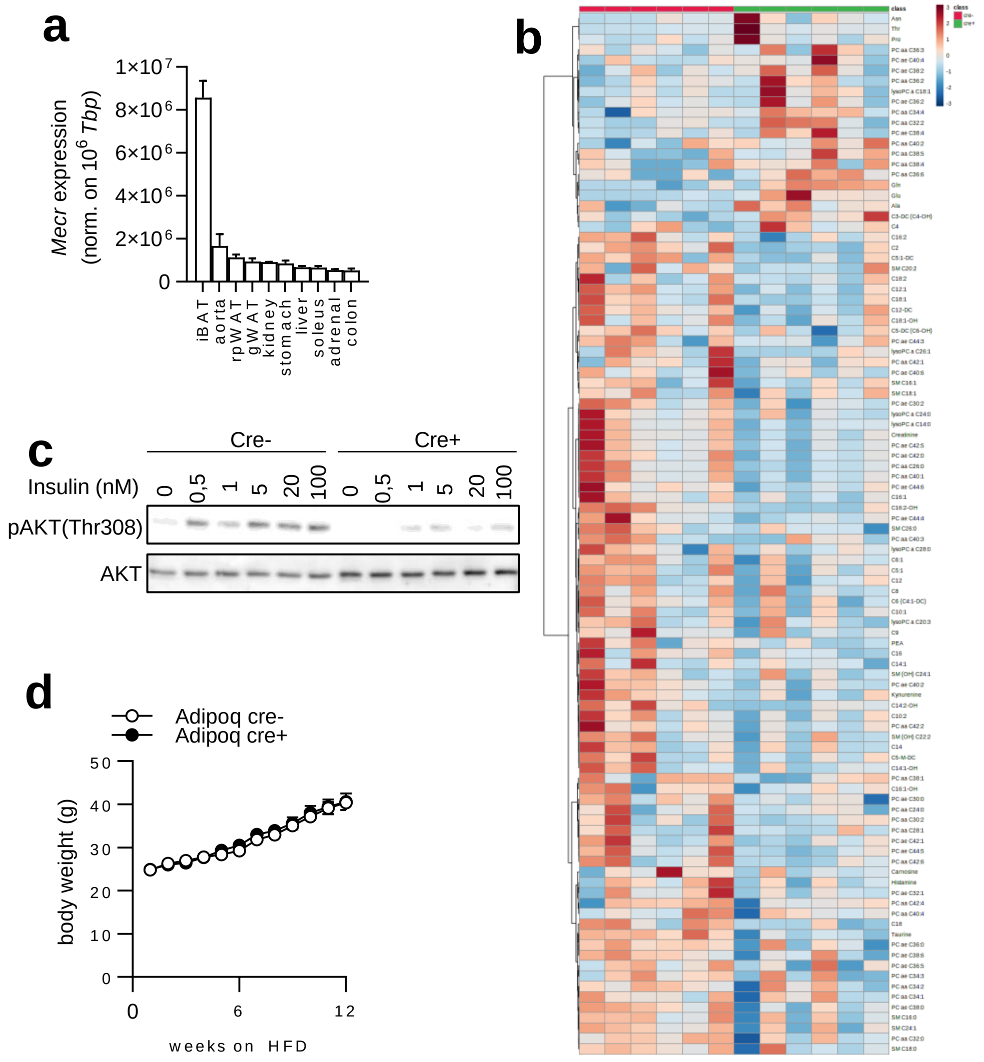


**Fig.S1:** (a) Gene expression of C57Bl6/J mice (n=3) organ panel. (b) Heat map of top 100 metabolite differences in white adipose tissue in *Cre*$^{-}$ and *Cre+ Adipoq-Cre Mecr*$^{flox/flox}$ mice. (c) Western blot of primary white adipocytes treatment with indicated concentrations of Insulin. (d) Body weight, of *Mecr*$^{flox/flox}$ *Adipoq-Cre* mice (n=9-10). Error bars indicate standard error of the mean.

#### Fig.S2

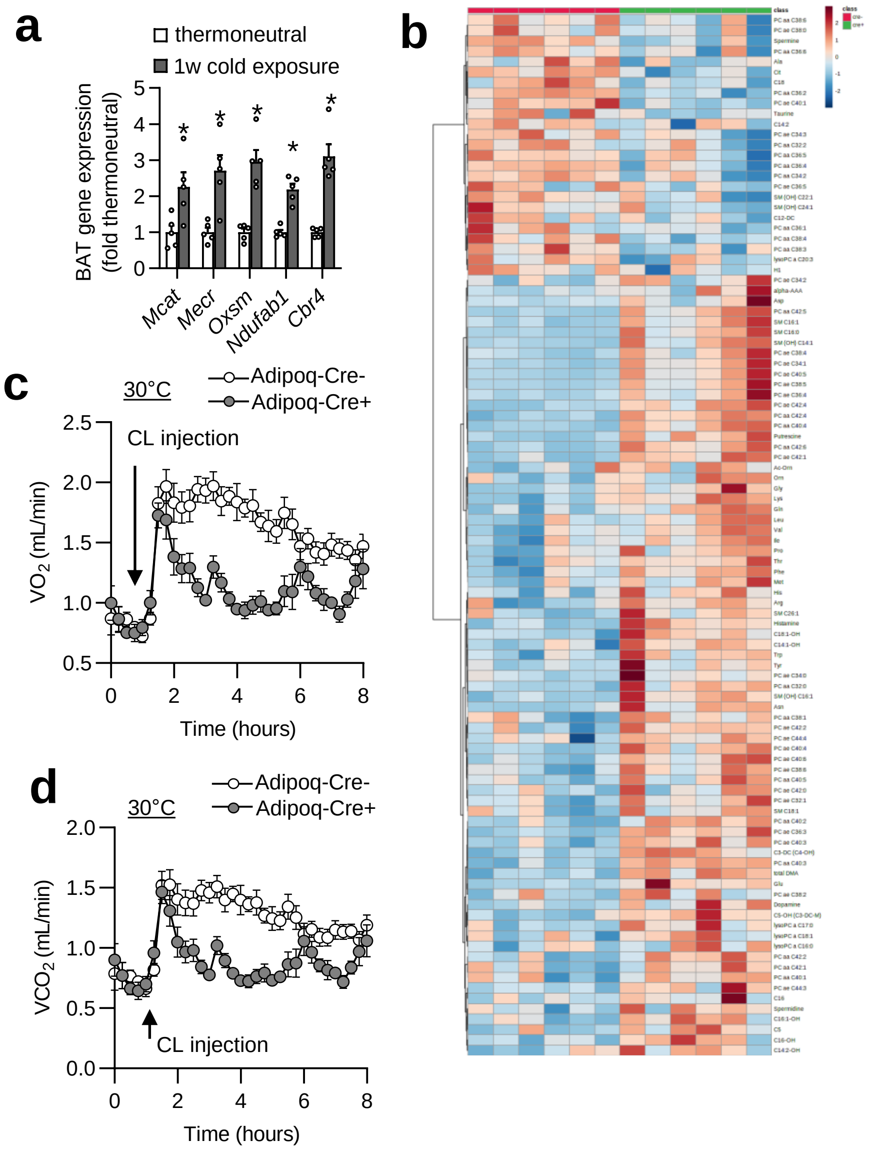


**Fig.S2:** (a) Gene expression of C57Bl6/J mice (n=3) organ panel. (a) Body weight, of Mecrflox/flox Adipoq-Cre mice (n=9-10). Thermoneutrality (1w) vs (1w) cold exposed wild type mice (n=5). (b) Heat map of top 100 metabolite differences in brown adipose tissue in *Cre*⁻ and *Cre+* *Adipoq-Cre Mecr*$^{flox/flox}$ mice. (c) oygen consumtption and (d) carbon dioxide production of *Mecr*$^{flox/flox}$ *Adipoq-Cre* mice injected with beta3-adrenergic agonist CL316,243 at thermoneutrality (n=6). Error bars indicate standard error of the mean. *=p<0.05 by Student's T-Test.

### Fig.S3

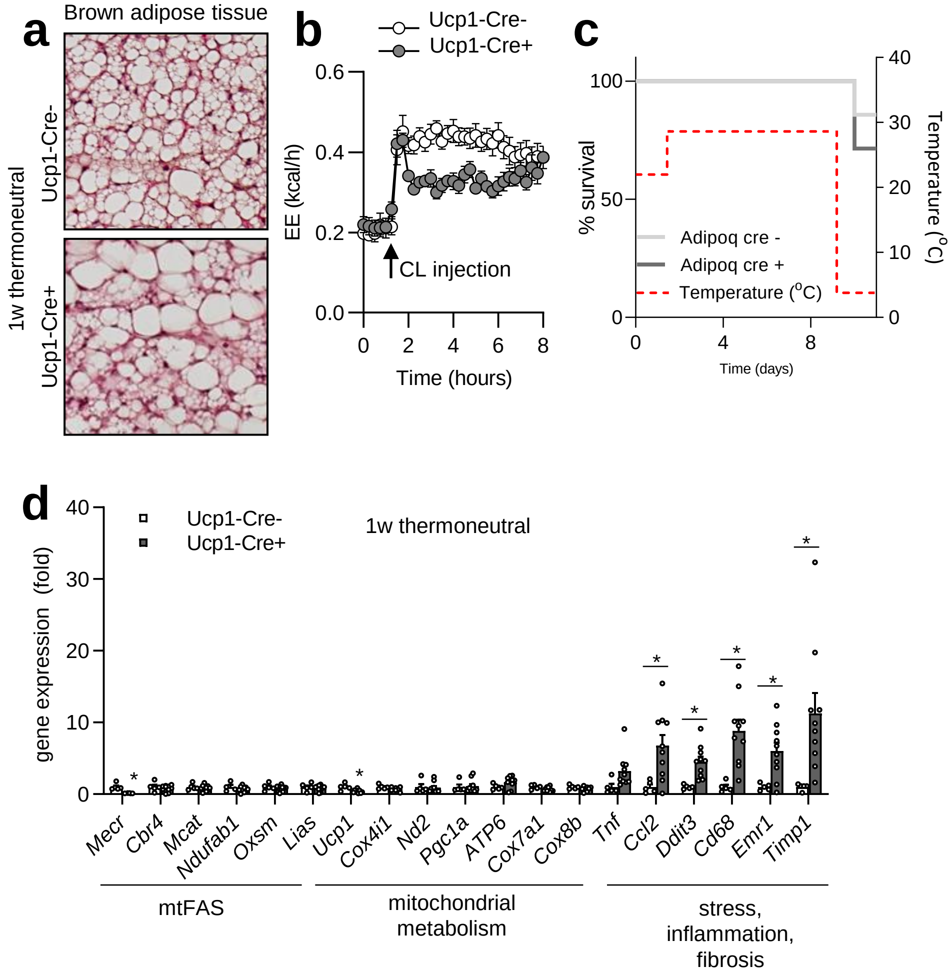


**Fig.S3:** (a) H&E staining or gene expression of BAT of *Mecr*$^{flox/flox}$ *Ucp1-Cre* mice (n=6-10). (b) Oxygen consumption of *Mecr*$^{flox/flox}$ *Adipoq-Cre* mice injected with beta3-adrenergic agonist CL316,243 at thermoneutrality (n=6). (c) Survival curve of *Mecr*$^{flox/flox}$ *Ucp1-Cre* mice housed at thermoneutrality and cold (n=6). Gene expression of BAT of *Mecr*$^{flox/flox}$ *Ucp1-Cre* mice (n=6-10). Error bars indicate standard error of the mean. *=p<0.05 by Student's T-Test.

### Fig.S4

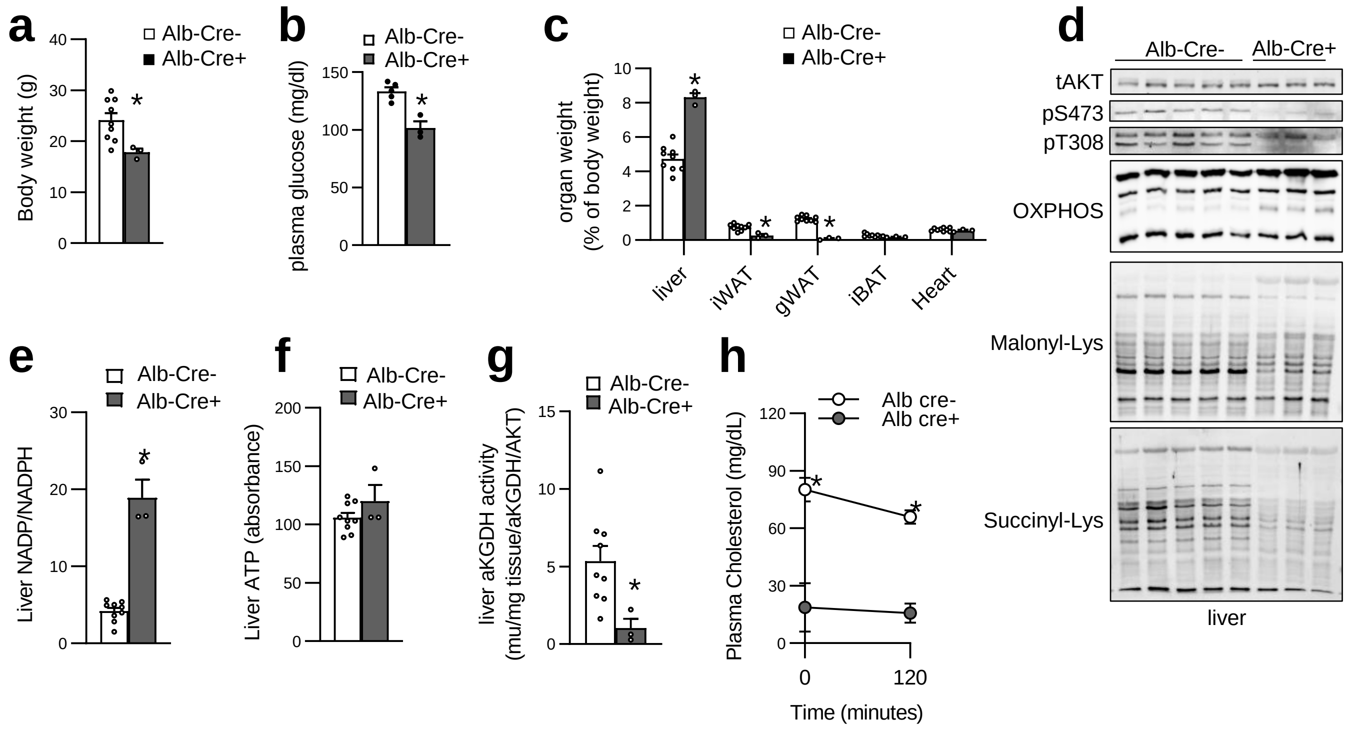


**Fig.S4:** (a) Body weight, (b) plasma glucose, (c) organ weights, (d) western blot, (e) NADP/NADPH ratio of livers, (f) ATP of liver, (g) plasma cholesterol and (h) plasma cholesterol of *Mecr*$^{flox/flox}$ *Alb-Cre* mice (n=3-9). Error bars indicate standard error of the mean. *=p<0.05 by Student's T-Test.

### Fig.S5

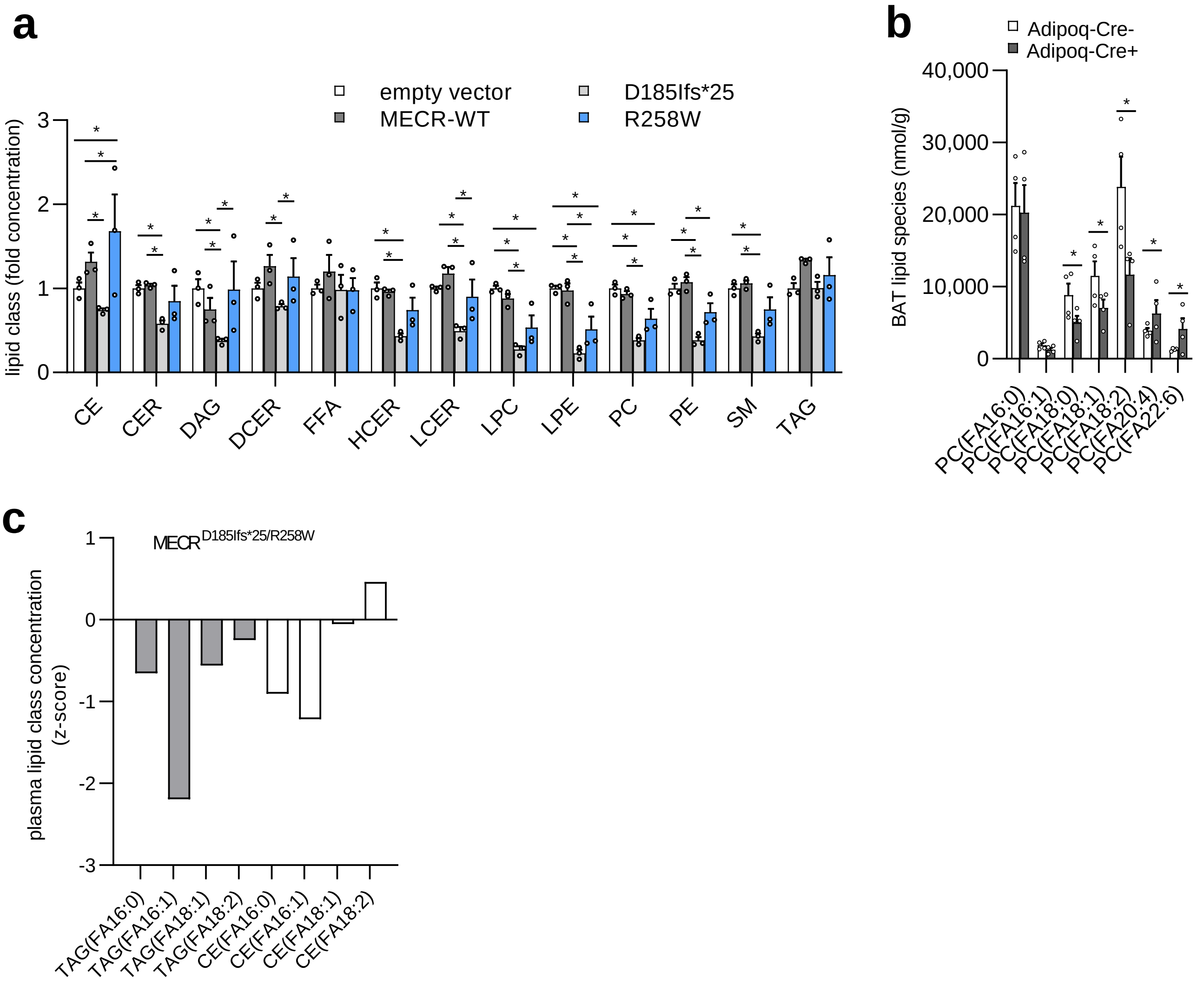


**Fig.S5:** LC/MS-based targeted lipidomics were performed from (a) HEK293T cells, which were overexpressed with either MECR wild type or indicated variants for 24h(n=3) or (b) brown adipose tissue from *Adipoq-Cre* mice or (c) plasma from the adult patient with indicated variants analyzed functionally in Fig.4 and FigS5a. Error bars indicate standard error of the mean. *=p<0.05 by Two Way ANOVA and Fishers LSD-Test.
